# BioLT: Streaming Biosignal Modeling

**DOI:** 10.64898/2026.11.06.26367429

**Authors:** Jinpei Han, Yuxuan Liu, Wei Tang, Louise Thwaites, David A. Clifton, Xiao Gu

## Abstract

Clinical biosignals such as ECG, PPG, and EEG are increasingly acquired as hours-long streams from bedside monitors and wearables, yet existing analysis pipelines almost universally operate on isolated short windows of around 10 seconds and cannot process the long, irregular, frequently interrupted signals. We introduce `BioLT`, a streaming decoder-only framework for long-context biosignal modeling that turns any window-level feature extractor or pretrained biosignal foundation model into a streaming model. `BioLT` combines (i) a discontinuity-aware encoding that exposes the inter-window elapsed time as an explicit input channel, so that the decoder can learn to model these gaps as clinically meaningful information, and (ii) a fixed-size compressed memory bank supervised with cumulative summary statistics, which keeps both training and inference linear in the recording length while preserving long-range context. We evaluate on CPSC-2021, VitalDB, MC-MED, and CHB-MIT for arrhythmia, sepsis, and seizure detection and for forecasting up to 20 minutes ahead, using domain features and embeddings from six biosignal foundation models. Across 21 dataset-feature settings, `BioLT` achieves the highest detection AUROC in 15 and the highest 20-minute forecasting AUROC in 13 settings, and shows consistent performance gains over state-of-the-art snapshot foundation-model embeddings. These highlight the importance of modeling how streaming biosignal representations evolve over time, beyond improving window-level representations alone.

## 1 Introduction

Human biosignals are continuous streams of physiological information. Signals such as electrocar-diography (ECG), photoplethysmography (PPG), and electroencephalography (EEG) are increasingly acquired through wearable sensors, bedside monitors, and ambulatory devices, yielding long-term streams of physiology-state information. Their “streaming” nature makes them particularly valuable for clinical monitoring, where models are expected to continuously update patient-state estimates, and facilitate the early detection or forecasting of health deterioration (Blythe et al., 2022; Hannun et al., 2019; Hyland et al., 2020; Han et al., 2025).

However, existing biosignal AI methods are limited in their capacity to support such streaming analysis, as they typically operate in either of the following two extremes, as shown in Figure 1b.

**Figure 1:**
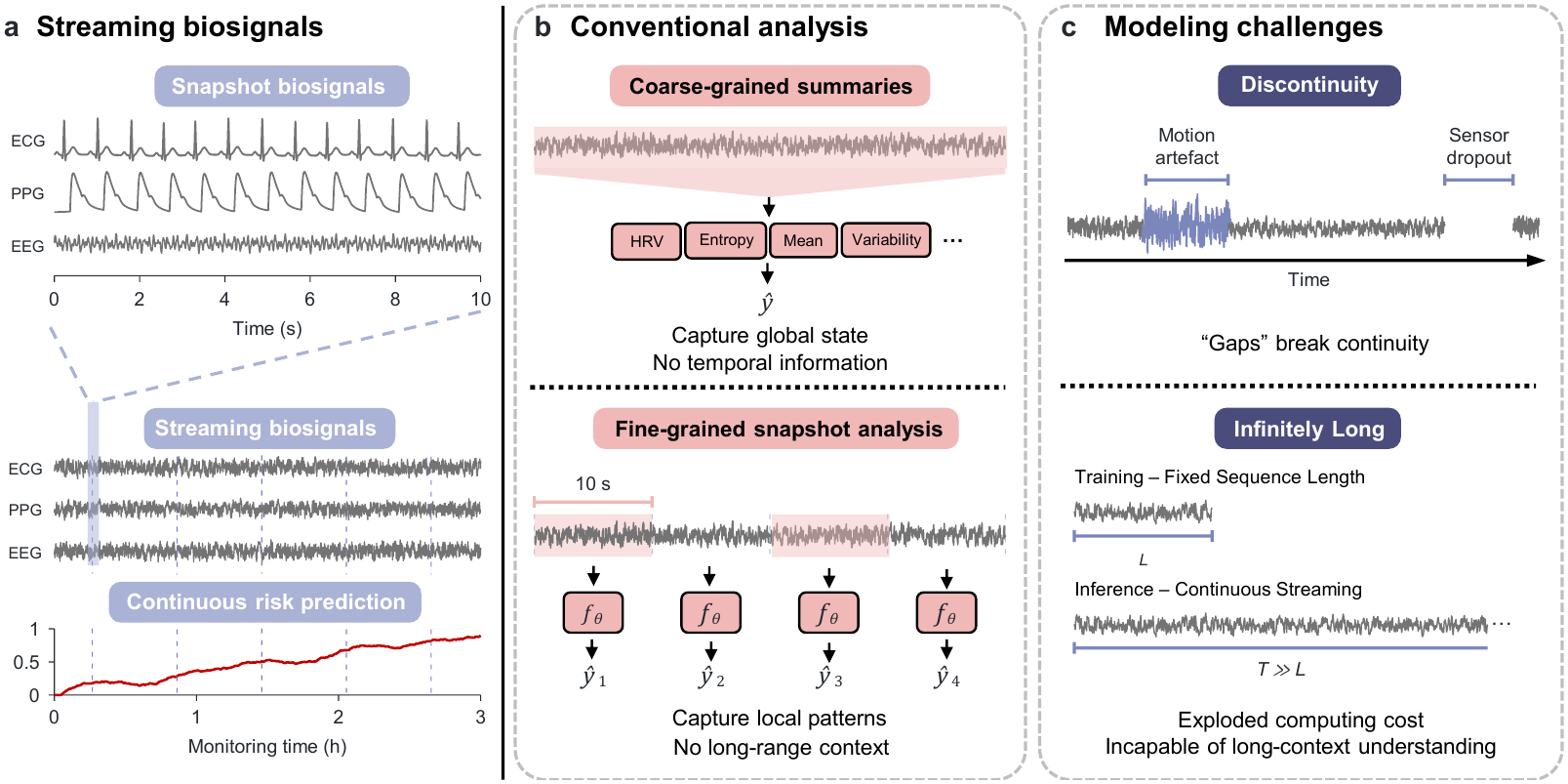
Challenges of streaming biosignal modeling. Illustration of two challenges of streaming biosignal modeling addressed by `BioLT`: the *discontinuity* of biosignal streams (gaps from missing or unreliable segments) and the *infinitely-long* nature of continuous monitoring (predictions must update as the trajectory unfolds), compared to coarse-grained summaries and fine-grained snapshot analysis.

1. *Coarse-grained summaries*, where summary statistics, such as heart rate variability and entropy distribution, are calculated from long-term duration biosignals (Duong et al., 2019; Hosseini et al., 2020). These features can capture global physiological characteristics, but they discard temporal ordering and therefore obscure how patient state unfolds over time.
2. *Fine-grained snapshot analysis*, which operates on short biosignal segments (typically 10 seconds, i.e., windows), to derive physiologically meaningful features. These features may be manually engineered using domain knowledge (Elgendi, 2012), or more recently obtained as embeddings from foundation models (Gu et al., 2025; Li et al., 2025; Shu et al., 2025; Gu et al., 2026; Saha et al., 2025; Jiang et al., 2024; Wang et al., 2025). While effective for local pattern recognition, such methods usually treat each window as an independent sample during training or evaluation, ignoring the continuous trajectory of the underlying physiology.

Several works attempt to bridge this gap by applying sequential models over window-level “snapshot” features. They either perform pre-hoc aggregation of these window-level features, or post-hoc aggregation of window-calculated probability/logits (Supratak et al., 2017; Phan et al., 2019; Tison et al., 2018). However, they mostly need to fix a maximum sequence length during training, and could not perform properly when the biosignal is streaming over the long term - the very practical needs necessitated by continuous monitoring.

To model streaming biosignals, there are two fundamental challenges inherent to such streaming nature, which need to be addressed properly, as shown in Figure 1c.

### (i) “Discontinuity” of streaming biosignals

One particular “sweet spot” of biosignals is their continuous nature - supporting continuous-monitoring needs. In reality, however, biosignal streams are rarely perfectly continuous. “Gaps” (no signals or unreliable segments) often arise during signal acquisition due to issues such as poor sensor contact or motion artifacts. In conventional window-level analysis, such low-quality windows may often be discarded. However, for streaming trajectory modeling, simply removing these windows breaks temporal continuity, especially for long missing intervals. Naive imputation is also insufficient, especially when gaps are long.

### (ii) “Infinitely-Long” nature of streaming biosignals

Biosignal streams can, in principle, continue indefinitely as new observations arrive over time. This creates a challenge for models that rely on fixed-length inputs or predefined observation windows. In continuous monitoring, the model should move beyond retrospective analysis of fixed-length segments and continuously update its predictions as the physiological trajectory evolves. This requires the model to preserve local waveform information while capturing long-range temporal dependencies across an expanding stream.

To address these challenges, we propose `BioLT` (**Bio**signal **L**ong-context **T**ransformer), a streaming framework that turns snapshot-level biosignal features or foundation-model embeddings into causal patient trajectories for real-time streaming prediction. `BioLT` combines two designs: (i) a discontinuity-aware encoding that exposes inter-window elapsed time to the model (Figure 2), and (ii) a compressed streaming “memory” that preserves long-range context into a fixed number of tokens, to avoid quadratic attention costs over long streaming biosignals (Figure 4). Across ECG, PPG, and EEG tasks spanning arrhythmia, sepsis, and seizure, for both detection and forecasting, `BioLT` improves over snapshot-level and temporal aggregation baselines, supporting its ability to model biosignals as long, irregular, continuously updated streams.

**Figure 2:**
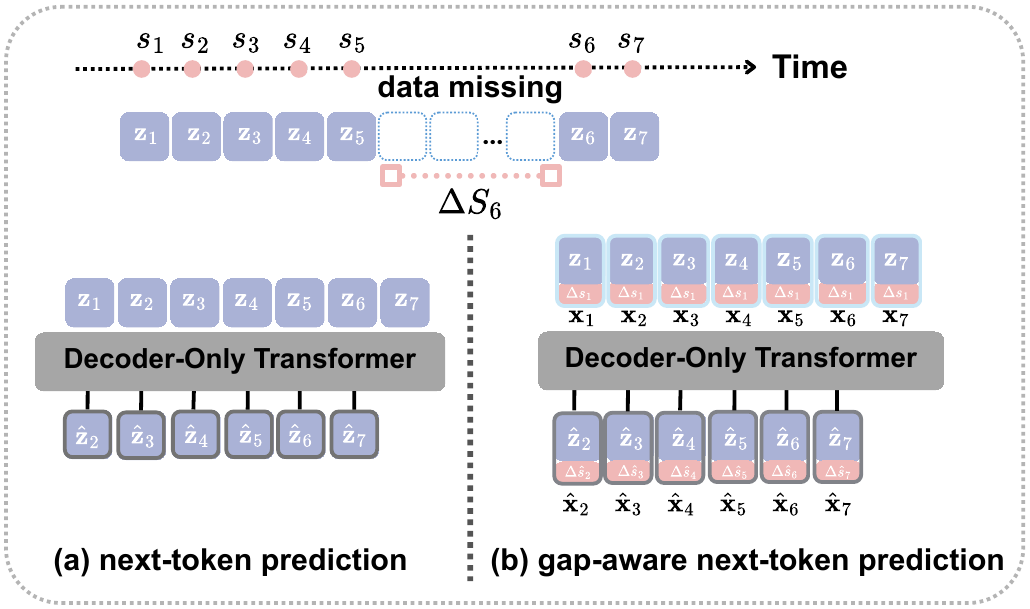
Discontinuity-aware token encoding. Neighboring valid biosignal windows may be separated by irregular elapsed time due to missing or low-quality signal segments. `BioLT` exposes this gap as Δ*s*_*t*_ in the input token.

## 2 Related Work

### Biosignal modeling

Biosignal analysis is evolving in terms of both how physiological representations are learned from raw signals, and how biosignal models are guided during learning (Hong et al., 2020; Gu et al., 2025). Early approaches relied heavily on domain-driven feature engineering (such as morphological features, frequency-domain measures, and signal-quality indicators) from raw biosignals (Faust et al., 2018; Gu et al., 2025). They typically need sophisticated signal processing and a feature selection procedure, necessitating substantial expert design. Deep learning methods have shifted such analysis from handcrafted features to learn task-relevant embeddings via careful annotation (Hannun et al., 2019; Supratak et al., 2017; Tison et al., 2018), and more recently to foundation models supporting general-purpose representations by self-supervised learning (Li et al., 2025; Shu et al., 2025; Gu et al., 2026; Saha et al., 2025; Jiang et al., 2024; Wang et al., 2025). However, most of these methods still focus on short, fixed-length windows, treating biosignals primarily as isolated snapshots rather than continuous physiological streams, leaving long-context modeling and continuous-monitoring evaluation underexplored for biosignals.

### Sequence modeling

Sequential modeling has been widely studied across language, video, and time series, with recurrent networks, Transformers, and state-space models providing common tools for modeling these sequential data. A central challenge is how to extend models beyond short contexts. Existing methods such as architecture variants (Rae et al., 2020), attention variants (Press et al., 2022), and state-space models (Lahoti et al., 2026) aim to improve memory, extrapolation, or computational efficiency for long sequences. A complementary direction is to carry history in a “memory” module, which caches hidden states from previous token attentions (Dai et al., 2019). These concepts are relevant to our problem, where monitoring records may extend over long periods. However, biosignal streams introduce additional practical challenges: valid windows may be interrupted by missing or unreliable recordings, and the elapsed time between neighboring windows can itself be informative.

### Real-time clinical prediction

Real-time clinical prediction aims to update risk estimates as new patient data arrive. This has been implemented using electronic health record (EHR)-based models, such as early warning scores (e.g., the Sequential Organ Failure Assessment (SOFA) or the National Early Warning Score (NEWS)) based on intermittently observed vital-sign and laboratory data (Reyna et al., 2020; Hyland et al., 2020). In contrast, biosignal-based systems, due to their “continuity”, are better suited for generating real-time risks of deterioration or adverse events (Hannun et al., 2019; Tison et al., 2018). Nevertheless, in both settings, models are commonly evaluated at the window level or within a fixed-length temporal context. This leaves open the question of how to model and evaluate risk trajectories over long, irregular, continuously updated biosignal streams.

## 3 Method

Given streaming biosignals, we propose `BioLT` as a framework for modeling continuous physiological trajectories and supporting real-time risk prediction. `BioLT` builds on snapshot-level biosignal representations. Given a raw biosignal recording, we first divide it into short valid non-overlapping windows (in practice, 10 seconds) and extract one representation for each window, using either hand-crafted physiological features or embeddings from a pretrained biosignal foundation model. `BioLT` then models the resulting sequence as a causal physiological trajectory.

Let **z**_*t*_ ∈ ℝ^*D*^ be the representation of the *t*-th valid window, where *D* denotes the representation dimension, and let *s*_*t*_ be its timestamp on the clinical timeline. A recording is represented as

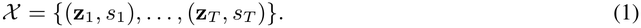

At each valid window *t*, `BioLT` produces *ŷ*_*t*_ = *f*_*θ*_(**z** ≤ _*t*_, *s* ≤ _*t*_), where *ŷ*_*t*_ denotes the predicted risk or class probability, and *y*_*t*_ denotes the corresponding task-specific label for detection (whether an adverse event happens at the current timepoint) or forecasting (whether adverse event will happen in the future). The prediction is causal: *ŷ*_*t*_ depends only on observations available up to timestamp *s*_*t*_.

`BioLT` uses a decoder-only temporal backbone to update the patient representation under a causal attention mask (Section 3.1). To address the practical challenges of streaming biosignals, we introduce two components: a discontinuity-aware token encoding that exposes inter-window elapsed time to the model (Section 3.2), and a compressed memory bank that preserves long-range trajectory context while keeping inference efficient (Section 3.3).

### 3.1 Basic Modeling: Decoder-only next-token prediction

For real-time monitoring, predictions must be made using only past and current information. This aligns the standard causal sequence-modeling setting, which has been widely used in language (Brown et al., 2020) and video (Zhao et al., 2023; Girdhar & Grauman, 2021) modeling. `BioLT` therefore uses a decoder-only temporal backbone that processes the window sequence from left to right. Given the input tokens **z**_1:*T*_, the decoder produces hidden states **h**_1:*T*_ = *F*_*θ*_(**z**_1:*T*_ ), where a causal attention mask ensures that **h**_*t*_ depends only on **z**_≤*t*_. **h**_*t*_ is then used for prediction at timestamp *s*_*t*_.

To learn from biosignal streams, we pretrain `BioLT` with a next-window prediction (i.e., autoregressive) task. For two neighboring windows, let Δ**z**_*t*+1_ = **z**_*t*+1_ ™ **z**_*t*_ denote the change in their snapshot representations. `BioLT` predicts this change from current summary vector: 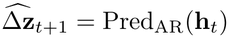, where Pred_AR_ is a lightweight prediction head. The pretraining loss is mean squared error between the predicted and true changes, averaged over neighboring valid window pairs:

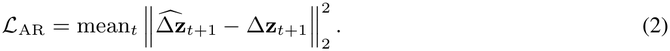

By predicting changes between windows, `BioLT` learns temporal physiological dynamics that are useful for downstream detection and forecasting.

### 3.2 Module 1: The “discontinuity” of streaming biosignals

The basic causal decoder provides the streaming backbone of `BioLT`. The next question is how to form input tokens for biosignal streams, which are often not perfectly continuous in practice, as mentioned before (due to factors such as noise and sensor drop-out). As a result, two neighboring valid windows may be separated by a long elapsed time (*s*_*t*_ ™ *s*_*t* ™ 1_). Direct imputation is often unreliable, especially when gaps are long. Conversely, treating these windows as equally spaced can make the model confuse missing time with continuous physiological change.

We take inspirations from sequential modeling of EHRs, where “irregularity” is a persistent issue: valid clinical records may be observed at different and unevenly spaced time points. In recent Transformer-based EHR models (Renc et al., 2024), such gap information is often treated not merely as missingness to be imputed, but as a clinically meaningful signal that may reflect patient state, clinical workflow, or monitoring intensity. Instead of forcing all observations onto a regular time grid, these models encode elapsed time between observations as part of the input.

Similar “informativeness” of gap is also noted in biosignal gaps, as demonstrated in Figure 3. Therefore, we adopt the same principle for biosignal streams: rather than filling long recording gaps, we make the elapsed time between neighboring valid windows visible to `BioLT`.

**Figure 3:**
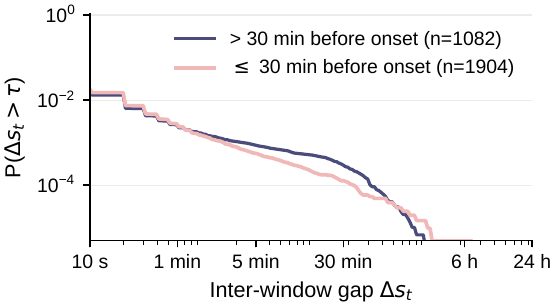
Inter-window gaps are “informative” to adverse events. Pooled CCDF of Δ*s*_*t*_ on MC-MED ECG, comparing windows within 30 minutes before sepsis onset with those outside the forecast horizon. The different gap distributions suggest that elapsed time provides useful context for streaming biosignal prediction.

To encode this information, we define the inter-window elapsed time as

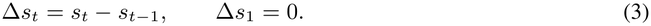

The model input token for the *t*-th valid window is then **x**_*t*_ = [**z**_*t*_; Δ*s*_*t*_]. The augmented token **x**_*t*_ is then used throughout `BioLT`, including in the next-window prediction objective of Section 3.1, as shown in Figure 2.

### 3.3 Module 2: The “Infinite” Streaming of biosignals

Apart from the “gap” issue, the other challenge is “infinite length” incurred by real-world streaming biosignals. Such streaming may continue for hours or days, producing an increasing number of valid windows. For instance, in our datasets, individual recordings reach up to 5,907 valid windows (∼16 h) on VitalDB. An ideal model should therefore update continuously, while not storing or attending to every previous window, to avoid prohibitive quadratic attention cost. `BioLT` addresses this with a compressed memory, as illustrated in Figure 4.

**Figure 4:**
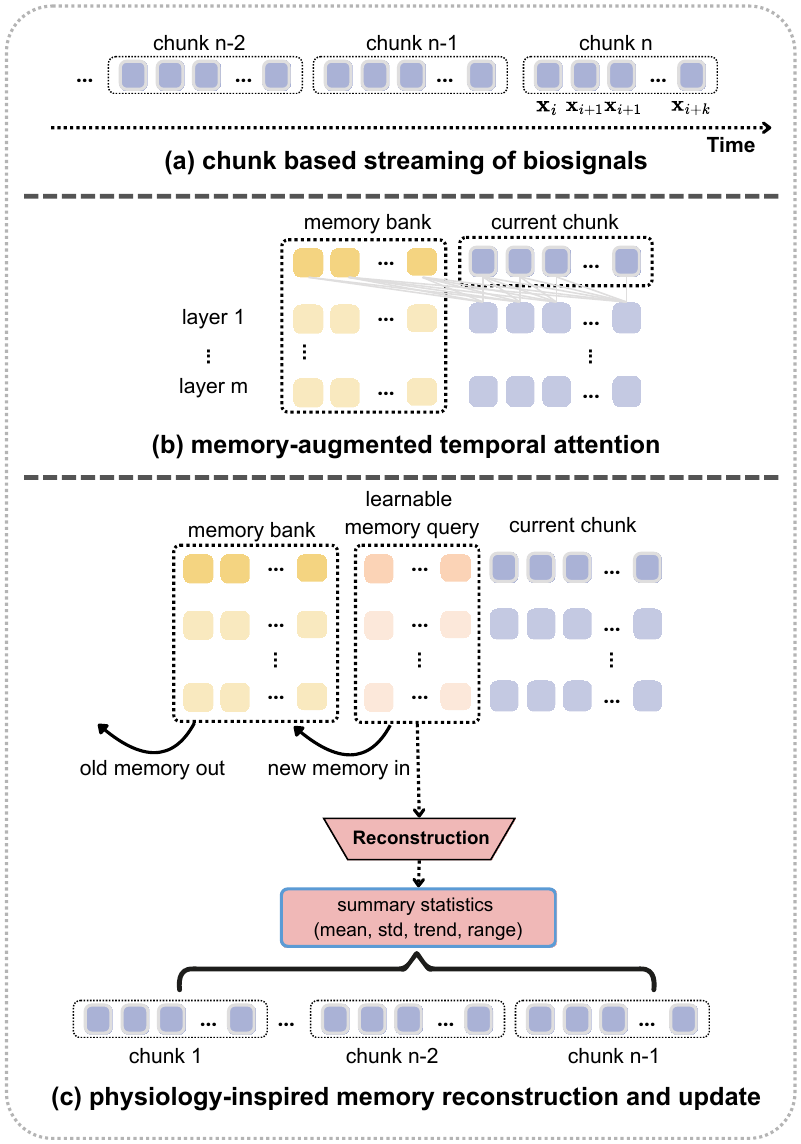
Compressed streaming memory in BioLT. **(a)** A long biosignal stream is divided into consecutive chunks. **(b)** Each temporal layer attends to both the current chunk and a fixed-size compressed memory bank. **(c)** Learnable memory queries produce new memory slots, by compressing the attention between old memory bank and last chunk; these slots are supervised by physiology-inspired trajectory summaries, and used to update the memory bank, in a first-in-first-out manner.

#### Chunk based streaming

Rather than attending to the whole sequence, we first divide the input into consecutive chunks comprising multiple windows (empirically set as 180 windows). Within the current chunk, `BioLT` applies exact causal attention for temporal modeling.

#### Memory-augmented temporal attention

To carry information across chunks, each temporal layer also attends to a fixed-size compressed “memory bank” *M*^cmp^ (Figure 4b), which stores compact summaries of previous chunks, and keeps updating as new chunks arrive, in a recurrent manner. Predictions therefore use both causal context within the current chunk and compressed information from earlier chunks.

#### Physiology-inspired memory learning

To make sure the “memory bank” preserves long-context historic information, we apply a learnable query module to compress the attentions operated between current chunk and old-”memory bank” into a small set of memory slots using learnable queries, and append these slots to the “memory bank” in a FIFO (first-in first-out) manner.

##### (i) Learnable query of current chunk

The memory compression is based on a learnable query mode. As shown in Figure 4c, after processing the current chunk, learnable memory query module compresses its hidden states into *K* ordered memory slots, **c**_*k*,1_, …, **c**_*k,K*_. These slots are added to *M*^cmp^, while the oldest slots are removed when the bank exceeds its capacity.

##### (ii) Learning objectives

To make sure the compressed memory preserves clinically meaningful information about the whole past trajectory, we regularize the learning using cumulative summary targets inspired by conventional long-term biosignal analysis. In conventional analysis, long recordings are often described by summary statistics such as average, variability, trend, and amplitude range.

Hence, we define **q** = [*µ, σ, b, r*], where *µ, σ, b*, and *r* denote the mean, standard deviation, linear trend, and dynamic range of the hidden states observed up to that point. A lightweight reconstruction head predicts these statistics from the compressed slot, giving *ℒ*_mem_ = mean SmoothL1 (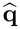, **q**), where 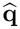 is the predicted summary target. The full pretraining objective is

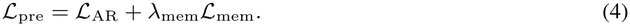

`BioLT` is trained in two stages, and in both stages, the memory bank is updated sequentially using truncated backpropagation through time (TBPTT), as in previous sequential modeling work (Mikolov et al., 2010). Each prediction conditions on long-range context while backpropagation stays local to the current chunk. During the pretraining stage, `BioLT` is optimized by Equation 4 on the whole, unlabeled recordings. After pretraining, `BioLT` is fine-tuned for downstream dense prediction by applying the task loss to the output at each valid window of labeled segments of at most 500 windows.

During inference, chunks are processed sequentially, the memory bank is updated after each chunk, and predictions are produced causally over the full recording.

## 4 Experimental Setup

### 4.1 Datasets

#### Biosignal datasets

We evaluate `BioLT` across ECG, PPG, and EEG recordings with diverse sampling rates, label structures, and clinical time scales. (i) **CPSC-2021** (Wang et al., 2021) contains dynamic ECG recordings for atrial fibrillation (AF) analysis; each window is labeled as non-AF, paroxysmal AF, or persistent AF. (ii) **VitalDB** (Eun et al., 2026) provides intraoperative ECG and PPG waveforms with rhythm annotations, indicating AF or not. (iii) **MC-MED** (Kansal et al., 2025) contains visit-level ECG and PPG waveforms for sepsis onset modeling, evaluated as binary event detection and forecasting. **(iv) CHB-MIT** (Shoeb, 2009) contains pediatric scalp EEG recordings with seizure onset annotations. These datasets contain dense annotations of those specific adverse events, with details summarized in Table 1.

**Table 1:** Dataset summary. Evaluation covers 3 modalities and 4 datasets; records and 10-second windows are reported after preprocessing.

| **Dataset** | **Mod.** | **Task** | **Labels** | **Rec.** | **Win.** |
|---|---|---|---|---|---|
| CPSC-2021 | ECG | AF | 3-class | 1,424 | 170k |
| VitalDB | ECG | AF | Binary | 431 | 571k |
| VitalDB | PPG | AF | Binary | 382 | 470k |
| MC-MED | ECG | Sepsis | Binary | 2,510 | 1.65M |
| MC-MED | PPG | Sepsis | Binary | 2,715 | 2.12M |
| CHB-MIT | EEG | Seizure | Binary | 655 | 342k |

#### Preprocessing

Each dataset is converted into per-recording sequences of non-overlapping 10-second windows, where each window has a timestamp *s*_*t*_ and snapshot representation **z**_*t*_ (by either domain feature engineering or foundation model, as detailed in Section 4.2). Signal cleaning and feature extraction are performed before temporal modeling.

#### Split and Task Setup

All experiments use patient-level 60/20/20 train/validation/test splits, identical across all experiments, to ensure fair comparison and avoid cross-recording leakage. For **detection**, *y*_*t*_ denotes the clinical state at timestamp *s*_*t*_; for **forecasting**, 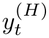 indicates whether the target event occurs within [*s*_*t*_, *s*_*t*_ + *H*]. We evaluate *H* ∈ {1, 5, 10, 20} minutes, with *H* = 0 treated as detection.

### 4.2 Compared Methods

#### Feature-wise

We consider both hand-crafted domain features and foundation-model-derived embeddings to represent each 10-second “snapshot” window. (i) For **ECG**, domain features cover morphology, heart-rate-variability, and signal-quality features (64 features); foundation-model embeddings are extracted with CSFM (Gu et al., 2026), ECGFounder (Li et al., 2025), and CLEF (Shu et al., 2025). (ii) For **PPG**, domain features cover morphology, and HRV indices derived from systolic-peak intervals, and signal-quality features (71 features); embeddings are extracted with CSFM (Gu et al., 2026) and PulsePPG (Saha et al., 2025). (iii) For **EEG**, per-channel domain features cover time-domain statistics, frequency-band powers, wavelet time-frequency descriptors, and complexity/entropy measures (31 features ×22 channels); foundation-model embeddings are extracted with LaBraM (Jiang et al., 2024) and CBraMod (Wang et al., 2025).

#### Temporal-modeling-wise

We compare with several temporal aggregation strategies. **Snapshot** applies an MLP classifier independently to each window and has no temporal context. **MeanPool** and **MaxPool** classify cumulative feature averages or maxima up to the current time. **LSTM** uses recurrent hidden states to carry history across windows. **TCN** applies causal dilated temporal convolutions. **Attention** is a single-head causal self-attention layer that learns a weighted sum of past projected snapshots, subsequently classified by an MLP head. Mamba-3 (Lahoti et al., 2026) is a recent state-space sequence model whose selective scan provides linear-time recurrent context aggregation. **Transformer-XL** (Dai et al., 2019) is architecturally identical to `BioLT`, whereas the compressed memory bank is replaced by an uncompressed 180-window FIFO segment memory, isolating the effect of compression itself. It is trained with the same input tokens, autoregressive pretraining recipe, downstream training procedures, as `BioLT`.

In practice, to mimic realistic “streaming” settings, all compared methods are trained on sequences cropped to a fixed maximum length of 500 windows during downstream training and tested on the full recordings. Results are reported as mean±standard deviation over three random seeds.

### 4.3 Performance Metrics

#### Window-based metrics

At each 10-second window *t*, each model outputs a probability *p*_*t*_ under the causal setting. We evaluate window-level predictions using AUROC and F1. AUROC is independent of any threshold choice. For multi-class tasks, we report macro-averaged AUROC and F1, obtained by class-argmax without any operating threshold; for binary tasks, we report standard binary AUROC and F1 (95% specificity), where the operating threshold is calibrated per dataset/model/seed to attain 95% specificity on the *validation* split and then frozen before the test stream is scored.

#### Alarm-based metric

Apart from window-level metrics to assess individual predictions, continuous monitoring also requires evaluating whether clinical events are detected at an acceptable false-alarm burden. Following discussions with clinicians, we therefore add one event-level metric for the binary tasks. An *event* is a ground-truth occurrence of the target condition, i.e., a consecutive sequence of windows whose label is positive (*y*_*t*_ = 1); a model raises an *alarm* at window *t* when its predicted probability *p*_*t*_ exceeds a threshold *θ*. An alarm is *false* if no event begins within the following 30 minutes; an event is *detected* if at least one alarm falls inside it. Sens@1FA/h is the fraction of test events detected at a threshold (*θ*). We choose (*θ*) on the validation set to yield at most one false alarm per hour and keep it fixed for testing:

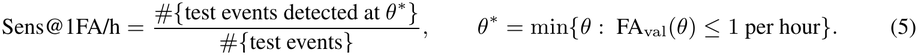

## 5 Results and Discussion

### 5.1 Main Detection and Forecasting Results

#### BioLT improves detection and forecasting across biosignal tasks, datasets, modalities, and feature types

Figure 5 provides a compact overview of the main results. The left panel summarizes detection performance at *H* = 0, using AUROC and F1 across ECG, PPG, and EEG tasks. The right panel summarizes forecasting performance averaged across *H* ∈ {1, 5, 10, 20} minutes. AUROC and F1 axes are normalized separately by the best observed value for each metric, respectively. A larger radius therefore indicates stronger relative performance. Overall, `BioLT` achieves the largest score on 8 of the 12 detection axes and on 8 of the 12 forecasting axes in Figure 5, including every forecasting AUROC axis. It is noteworthy that performance margins of forecasting is generally smaller, compared to detection, suggesting greater inherent difficulty of forecasting tasks.

**Figure 5:**
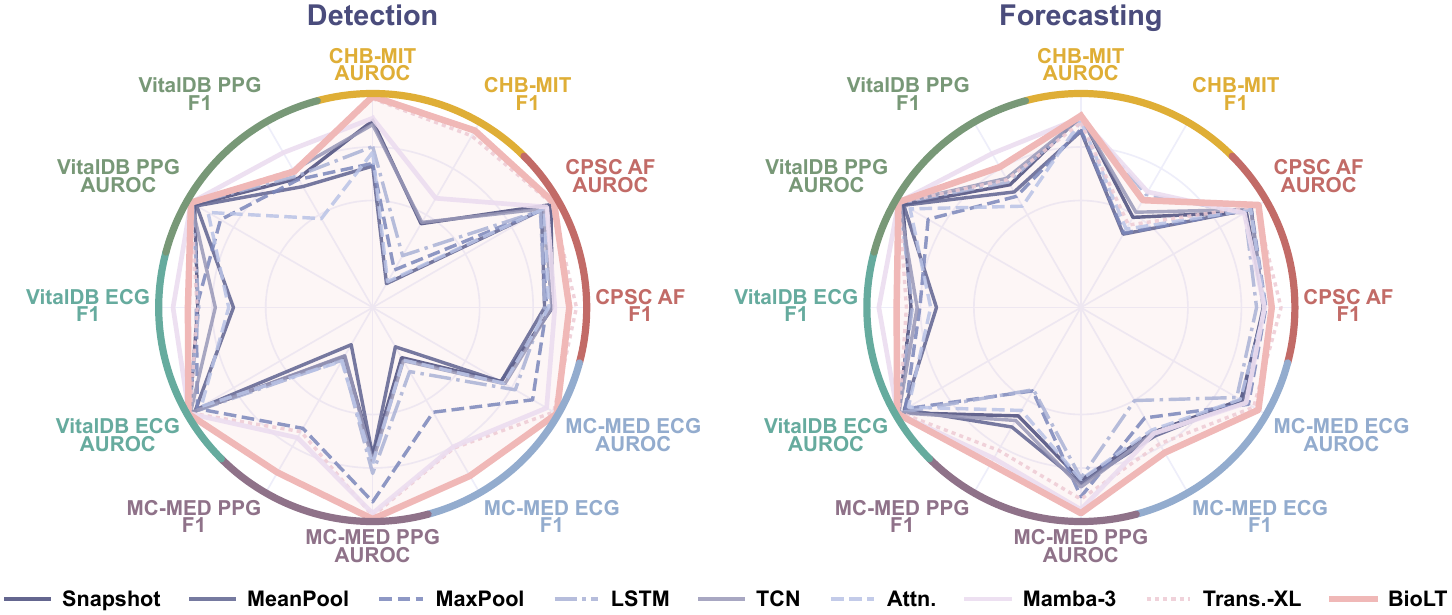
Hierarchical radar comparison of detection and forecasting performance (averaged across 1,5,10,20 min) across biosignal datasets. Each dataset is represented by two adjacent axes (AUROC and F1). The performance is averaged across all compatible features for each dataset.

#### BioLT is effective across both domain features and foundation-model embeddings

We further break down the results by dataset, compared method, and feature type in Table 2 for detection and Table 3 for 20-minute forecasting. Overall, `BioLT` achieves the best detection AUROC in 15 of 21 dataset-feature settings (Table 2) and the best 20-minute forecasting AUROC in 13 of 21 settings (Table 3). This demonstrates `BioLT` as an input-agnostic temporal backbone, compatible with both hand-crafted domain features as well as embeddings from pretrained foundation models.

**Table 2:** Detection results. Test AUROC (%, mean±std over 3 seeds). **Best mean** per row in **bold**, second best per row underlined. Rightmost column presents the performance gains using `BioLT` on domain features (Dom. `BioLT`) vs using snapshot on respective FM embeddings (Snap. FM).

| **Datasets** | **Features** | **Snapshot** | **MeanPool** | **MaxPool** | **LSTM** | **TCN** | **Attn.** | **Mamba-3** | **Trans.-XL** | **BioLT** | **Dom. BioLT**<br>**vs. Snap. FM** |
|---|---|---|---|---|---|---|---|---|---|---|---|
| **CHB-MIT**<br>EEG seizure | Domain | 75.85<sub>±3.60</sub> | 61.55<sub>±7.70</sub> | 54.82<sub>±7.95</sub> | 70.00<sub>±4.19</sub> | 69.92<sub>±3.32</sub> | 62.25<sub>±7.52</sub> | 76.15<sub>±4.45</sub> | 79.96<sub>±9.71</sub> | **81.97<sub>±7.89</sub>** |  |
|  | CBraMod | 69.89<sub>±5.36</sub> | 56.57<sub>±14.18</sub> | 53.24<sub>±1.94</sub> | 63.67<sub>±11.32</sub> | 69.50<sub>±9.16</sub> | 57.52<sub>±7.04</sub> | 74.40<sub>±6.23</sub> | **80.87<sub>±10.88</sub>** | 80.03<sub>±9.40</sub> | **+12.08** |
|  | LaBraM | 70.77<sub>±3.86</sub> | 45.82<sub>±4.98</sub> | 58.99<sub>±4.38</sub> | 52.87<sub>±14.51</sub> | 72.65<sub>±5.99</sub> | 60.30<sub>±2.70</sub> | 68.88<sub>±6.08</sub> | 81.34<sub>±3.41</sub> | **82.70<sub>±5.74</sub>** | **+11.20** |
| **CPSC-2021**<br>ECG AF | Domain | 92.81<sub>±2.58</sub> | 87.24<sub>±2.39</sub> | 86.02<sub>±3.41</sub> | 86.93<sub>±2.05</sub> | 88.28<sub>±4.21</sub> | 88.33<sub>±2.95</sub> | 91.58<sub>±2.50</sub> | 96.37<sub>±1.90</sub> | **96.83<sub>±1.69</sub>** |  |
|  | CSFM | 92.85<sub>±3.69</sub> | 88.02<sub>±6.26</sub> | 91.04<sub>±5.91</sub> | 89.85<sub>±4.06</sub> | 90.91<sub>±6.95</sub> | 88.87<sub>±8.05</sub> | 91.92<sub>±4.53</sub> | 93.32<sub>±5.30</sub> | **95.57<sub>±2.37</sub>** | **+3.98** |
|  | CLEF | **92.06<sub>±2.65</sub>** | 88.01<sub>±3.32</sub> | 82.78<sub>±7.16</sub> | 85.56<sub>±4.41</sub> | 86.23<sub>±6.18</sub> | 89.55<sub>±5.70</sub> | 88.06<sub>±4.54</sub> | 89.74<sub>±8.83</sub> | 90.23<sub>±4.57</sub> | **+4.77** |
|  | ECGFounder | 94.53<sub>±4.20</sub> | 89.43<sub>±4.94</sub> | 90.91<sub>±5.62</sub> | 88.20<sub>±7.35</sub> | 92.27<sub>±6.77</sub> | 91.56<sub>±6.35</sub> | 92.23<sub>±6.96</sub> | 95.73<sub>±3.36</sub> | **96.35<sub>±3.08</sub>** | **+2.30** |
| **VitalDB**<br>ECG AF | Domain | 92.84<sub>±1.36</sub> | 88.35<sub>±3.20</sub> | 84.79<sub>±1.98</sub> | 94.12<sub>±0.56</sub> | 91.81<sub>±1.64</sub> | 84.63<sub>±8.96</sub> | 94.47<sub>±0.86</sub> | 93.72<sub>±1.17</sub> | **94.91<sub>±1.58</sub>** |  |
|  | CSFM | 94.41<sub>±0.67</sub> | 91.35<sub>±2.15</sub> | 91.80<sub>±1.98</sub> | 94.56<sub>±0.17</sub> | 93.95<sub>±0.79</sub> | 91.11<sub>±3.16</sub> | 94.15<sub>±1.09</sub> | 93.92<sub>±0.67</sub> | **94.70<sub>±0.56</sub>** | **+0.50** |
|  | CLEF | 93.96<sub>±0.32</sub> | 91.16<sub>±0.91</sub> | 90.94<sub>±0.58</sub> | 93.40<sub>±0.23</sub> | 93.50<sub>±0.13</sub> | 86.71<sub>±3.63</sub> | 93.32<sub>±1.01</sub> | 93.44<sub>±0.89</sub> | **94.36<sub>±0.46</sub>** | **+0.95** |
|  | ECGFounder | 94.00<sub>±0.72</sub> | 91.92<sub>±1.99</sub> | 91.57<sub>±0.83</sub> | **94.43<sub>±0.72</sub>** | 94.08<sub>±0.69</sub> | 88.83<sub>±1.58</sub> | 94.30<sub>±1.43</sub> | 93.01<sub>±2.33</sub> | 93.48<sub>±0.52</sub> | **+0.91** |
| **VitalDB**<br>PPG AF | Domain | 88.90<sub>±0.48</sub> | 88.55<sub>±0.52</sub> | 79.07<sub>±1.92</sub> | 90.73<sub>±0.37</sub> | 90.54<sub>±0.47</sub> | 83.77<sub>±3.29</sub> | **91.98<sub>±0.47</sub>** | 90.83<sub>±0.58</sub> | 90.88<sub>±0.30</sub> |  |
|  | CSFM | 89.84<sub>±0.45</sub> | 88.28<sub>±0.85</sub> | 90.51<sub>±1.03</sub> | 90.97<sub>±0.51</sub> | 90.94<sub>±0.75</sub> | 87.27<sub>±2.08</sub> | 89.92<sub>±0.44</sub> | **91.13<sub>±0.30</sub>** | 90.65<sub>±0.73</sub> | +1.04 |
|  | PulsePPG | 86.37<sub>±0.15</sub> | 85.30<sub>±0.41</sub> | 56.92<sub>±3.76</sub> | 86.69<sub>±1.38</sub> | 88.63<sub>±0.32</sub> | 74.08<sub>±5.07</sub> | 88.61<sub>±0.65</sub> | **89.19<sub>±0.19</sub>** | 88.96<sub>±0.58</sub> | +4.51 |
| **MC-MED**<br>ECG sepsis | Domain | 54.23<sub>±0.96</sub> | 52.95<sub>±2.87</sub> | 62.76<sub>±2.87</sub> | 61.05<sub>±2.26</sub> | 57.31<sub>±2.54</sub> | 53.18<sub>±2.49</sub> | 74.37<sub>±2.10</sub> | 78.32<sub>±1.41</sub> | **78.78<sub>±0.63</sub>** |  |
|  | CSFM | 56.38<sub>±0.97</sub> | 57.29<sub>±1.72</sub> | 70.02<sub>±1.63</sub> | 62.49<sub>±0.92</sub> | 58.54<sub>±0.71</sub> | 58.50<sub>±3.11</sub> | 75.16<sub>±0.66</sub> | 77.71<sub>±0.88</sub> | **78.70<sub>±1.33</sub>** | **+22.40** |
|  | CLEF | 54.54<sub>±2.58</sub> | 55.13<sub>±2.77</sub> | 69.52<sub>±1.05</sub> | 63.63<sub>±3.25</sub> | 56.31<sub>±1.45</sub> | 57.67<sub>±3.60</sub> | 75.68<sub>±1.38</sub> | 78.89<sub>±1.12</sub> | **79.40<sub>±0.89</sub>** | **+24.24** |
|  | ECGFounder | 54.58<sub>±1.50</sub> | 54.61<sub>±1.12</sub> | 71.67<sub>±0.64</sub> | 57.22<sub>±4.24</sub> | 54.23<sub>±1.27</sub> | 55.75<sub>±1.94</sub> | 72.85<sub>±0.11</sub> | 75.44<sub>±0.49</sub> | **78.41<sub>±0.73</sub>** | **+24.20** |
| **MC-MED**<br>PPG sepsis | Domain | 55.54<sub>±0.53</sub> | 54.09<sub>±0.94</sub> | 73.78<sub>±0.90</sub> | 59.72<sub>±2.23</sub> | 58.51<sub>±1.12</sub> | 57.05<sub>±2.51</sub> | 78.42<sub>±0.58</sub> | 79.34<sub>±0.53</sub> | **80.96<sub>±1.10</sub>** |  |
|  | CSFM | 56.93<sub>±2.29</sub> | 56.78<sub>±3.53</sub> | 75.23<sub>±1.88</sub> | 66.33<sub>±2.51</sub> | 58.30<sub>±2.83</sub> | 60.44<sub>±2.30</sub> | 76.54<sub>±1.04</sub> | 78.82<sub>±0.64</sub> | **78.96<sub>±1.18</sub>** | **+24.03** |
|  | PulsePPG | 55.92<sub>±0.81</sub> | 57.64<sub>±2.31</sub> | 71.55<sub>±1.85</sub> | 62.25<sub>±1.26</sub> | 58.09<sub>±1.07</sub> | 60.06<sub>±0.87</sub> | 78.63<sub>±0.64</sub> | 78.35<sub>±1.26</sub> | **80.22<sub>±1.09</sub>** | **+25.04** |

**Table 3:** Forecasting results at horizon *H*=20 minutes. Test AUROC (%, mean±std over 3 seeds). **Best mean** per row in **bold**, second best per row underlined. Rightmost column presents the performance gains using `BioLT` on domain features (Dom. `BioLT`) vs using snapshot on respective FM embeddings (Snap. FM).

| Datasets | Features | Snapshot | MeanPool | MaxPool | LSTM | TCN | Attn. | Mamba-3 | Trans.-XL | BioLT | Dom. BioLT<br>vs. Snap. FM |
|---|---|---|---|---|---|---|---|---|---|---|---|
| **CHB-MIT**<br>EEG seizure | Domain | **60.04<sub>±8.08</sub>** | 57.49<sub>±10.33</sub> | 52.34<sub>±4.18</sub> | 58.89<sub>±6.90</sub> | 56.93<sub>±9.50</sub> | 56.86<sub>±16.89</sub> | 52.80<sub>±3.94</sub> | 52.43<sub>±9.90</sub> | 51.10<sub>±9.53</sub> | ↴ |
|  | CBraMod | 57.71<sub>±10.42</sub> | 58.18<sub>±16.99</sub> | 52.95<sub>±10.75</sub> | **60.51<sub>±13.95</sub>** | 58.88<sub>±13.07</sub> | 59.01<sub>±14.34</sub> | 56.24<sub>±9.36</sub> | 54.24<sub>±9.48</sub> | 56.62<sub>±13.16</sub> | **−6.61** |
|  | LaBraM | 47.83<sub>±5.86</sub> | 45.44<sub>±7.17</sub> | 53.90<sub>±12.04</sub> | 52.79<sub>±9.53</sub> | 54.45<sub>±12.07</sub> | 54.72<sub>±9.52</sub> | 52.86<sub>±6.85</sub> | 48.32<sub>±7.49</sub> | **56.69<sub>±13.83</sub>** | **+3.27** |
| **CPSC-2021**<br>ECG AF | Domain | 75.36<sub>±6.18</sub> | 78.53<sub>±9.37</sub> | 71.41<sub>±9.60</sub> | 77.53<sub>±5.01</sub> | 78.18<sub>±5.98</sub> | 81.94<sub>±3.39</sub> | 76.03<sub>±6.39</sub> | **87.11<sub>±2.76</sub>** | 86.43<sub>±1.63</sub> | ↴ |
|  | CSFM | 81.10<sub>±4.59</sub> | 83.47<sub>±1.90</sub> | 79.14<sub>±9.86</sub> | 82.77<sub>±4.97</sub> | 84.11<sub>±1.55</sub> | 81.40<sub>±3.09</sub> | 81.39<sub>±4.25</sub> | 82.66<sub>±2.09</sub> | **87.45<sub>±2.08</sub>** | **+5.33** |
|  | CLEF | 76.41<sub>±5.22</sub> | 80.30<sub>±4.80</sub> | 79.07<sub>±5.71</sub> | 77.91<sub>±6.04</sub> | 80.58<sub>±6.38</sub> | **82.02<sub>±7.21</sub>** | 79.14<sub>±5.97</sub> | 75.46<sub>±12.61</sub> | 79.67<sub>±9.31</sub> | **+10.02** |
|  | ECGFounder | 80.41<sub>±6.24</sub> | 79.33<sub>±4.43</sub> | 82.85<sub>±4.74</sub> | 75.27<sub>±5.08</sub> | 81.63<sub>±2.08</sub> | 79.99<sub>±2.40</sub> | 73.03<sub>±0.42</sub> | 83.64<sub>±2.29</sub> | **85.40<sub>±4.38</sub>** | **+6.02** |
| VitalDB<br>ECG AF | Domain | 88.75<sub>±2.12</sub> | 84.34<sub>±3.69</sub> | 82.02<sub>±3.68</sub> | 90.28<sub>±1.46</sub> | 88.97<sub>±2.08</sub> | 80.07<sub>±8.56</sub> | 90.67<sub>±1.28</sub> | 88.99<sub>±1.81</sub> | **91.34<sub>±1.53</sub>** | ↴ |
|  | CSFM | 89.40<sub>±1.79</sub> | 87.46<sub>±1.87</sub> | 87.81<sub>±0.56</sub> | 89.28<sub>±1.41</sub> | 89.74<sub>±1.44</sub> | 87.90<sub>±2.17</sub> | 90.09<sub>±1.51</sub> | 90.17<sub>±1.99</sub> | **90.61<sub>±1.51</sub>** | **+1.94** |
|  | CLEF | **89.35<sub>±1.75</sub>** | 86.17<sub>±1.09</sub> | 86.42<sub>±1.42</sub> | 88.55<sub>±2.30</sub> | 89.19<sub>±1.77</sub> | 86.13<sub>±2.77</sub> | 89.14<sub>±2.05</sub> | 88.24<sub>±2.78</sub> | 89.27<sub>±1.40</sub> | **+1.99** |
|  | ECGFounder | 89.57<sub>±1.52</sub> | 86.40<sub>±3.76</sub> | 87.78<sub>±1.48</sub> | 88.87<sub>±2.48</sub> | 90.12<sub>±1.68</sub> | 85.09<sub>±2.66</sub> | **90.32<sub>±1.06</sub>** | 89.02<sub>±2.80</sub> | 90.18<sub>±1.98</sub> | **+1.77** |
| VitalDB<br>PPG AF | Domain | 87.49<sub>±1.23</sub> | 88.36<sub>±1.54</sub> | 79.14<sub>±0.62</sub> | 88.81<sub>±1.26</sub> | 88.58<sub>±1.45</sub> | 87.49<sub>±2.42</sub> | 90.38<sub>±1.37</sub> | 89.19<sub>±1.55</sub> | **90.43<sub>±1.20</sub>** | ↴ |
|  | CSFM | 88.73<sub>±0.73</sub> | 88.68<sub>±0.64</sub> | 89.32<sub>±1.85</sub> | 88.94<sub>±0.56</sub> | 89.48<sub>±0.44</sub> | 87.89<sub>±0.28</sub> | 89.03<sub>±0.94</sub> | **89.81<sub>±0.96</sub>** | **89.81<sub>±1.08</sub>** | **+1.70** |
|  | PulsePPG | 85.05<sub>±1.52</sub> | 84.59<sub>±2.44</sub> | 51.36<sub>±9.28</sub> | 87.38<sub>±2.63</sub> | **88.38<sub>±1.09</sub>** | 81.64<sub>±9.41</sub> | 88.07<sub>±1.09</sub> | 87.48<sub>±1.22</sub> | 88.22<sub>±1.42</sub> | **+5.38** |
| MC-MED<br>ECG sepsis | Domain | 52.47<sub>±1.76</sub> | 53.26<sub>±2.44</sub> | 57.25<sub>±0.93</sub> | 52.84<sub>±1.01</sub> | 54.04<sub>±1.71</sub> | 54.18<sub>±1.37</sub> | 55.19<sub>±0.59</sub> | **63.24<sub>±2.16</sub>** | 62.20<sub>±1.10</sub> | ↴ |
|  | CSFM | 54.37<sub>±1.87</sub> | 53.94<sub>±0.93</sub> | 60.30<sub>±1.21</sub> | 53.42<sub>±3.18</sub> | 54.70<sub>±1.71</sub> | 51.71<sub>±3.41</sub> | 53.73<sub>±1.29</sub> | 55.06<sub>±0.99</sub> | **61.01<sub>±2.28</sub>** | **+7.83** |
|  | CLEF | 54.03<sub>±2.38</sub> | 55.10<sub>±3.96</sub> | 59.20<sub>±2.25</sub> | 54.26<sub>±1.00</sub> | 54.64<sub>±2.31</sub> | 55.48<sub>±3.42</sub> | 56.86<sub>±1.06</sub> | 54.72<sub>±2.08</sub> | **62.24<sub>±1.67</sub>** | **+8.17** |
|  | ECGFounder | 52.52<sub>±2.30</sub> | 52.19<sub>±1.87</sub> | 55.44<sub>±1.51</sub> | 52.00<sub>±1.69</sub> | 53.38<sub>±0.92</sub> | 52.81<sub>±1.91</sub> | 51.46<sub>±2.28</sub> | 55.01<sub>±2.34</sub> | **58.05<sub>±1.70</sub>** | **+9.68** |
| MC-MED<br>PPG sepsis | Domain | 52.27<sub>±0.73</sub> | 53.82<sub>±2.23</sub> | 60.20<sub>±0.82</sub> | 52.04<sub>±1.52</sub> | 54.27<sub>±0.79</sub> | 51.64<sub>±1.17</sub> | 62.03<sub>±0.56</sub> | 64.73<sub>±0.79</sub> | **64.92<sub>±0.71</sub>** | ↴ |
|  | CSFM | 52.34<sub>±1.56</sub> | 51.93<sub>±2.93</sub> | 60.81<sub>±1.75</sub> | 52.76<sub>±1.06</sub> | 53.56<sub>±2.15</sub> | 51.50<sub>±3.46</sub> | 58.93<sub>±2.40</sub> | 52.65<sub>±0.95</sub> | **63.15<sub>±0.58</sub>** | **+12.58** |
|  | PulsePPG | 51.19<sub>±0.48</sub> | 51.42<sub>±1.17</sub> | 60.14<sub>±0.64</sub> | 51.52<sub>±1.32</sub> | 52.77<sub>±0.46</sub> | 50.67<sub>±1.22</sub> | 61.66<sub>±1.47</sub> | 53.17<sub>±0.52</sub> | **63.88<sub>±0.80</sub>** | **+13.73** |

#### BioLT performance gains reflect the value of temporal context

The advantage of `BioLT` is most evident in settings where temporal context provides information beyond an isolated 10-second window. For instance, at detection (Table 2), `BioLT` performs particularly strongly on CHB-MIT seizure detection and MC-MED sepsis detection, while gains are smaller on AF tasks where local rhythm morphology is already informative. For forecasting, margins over competing methods are generally smaller. At 20-min forecasting (Table 3), however, `BioLT` remains strongest in most settings, particularly for MC-MED sepsis forecasting across both ECG and PPG, in line with prior evidence that dynamic physiology changes carry prognostic information in sepsis (Zhang et al., 2026).

#### Effective aggregation of domain features outperforms snapshot-based foundation model features

Further into detection results (Table 2), as shown in the rightmost column, `BioLT` on domain features outperforms the snapshot-based method on all FM embeddings of the same datasets. More-over, once modeled by `BioLT`, domain features and FM embeddings achieve broadly competitive performance across tasks, as in second-to-last column. This is particularly evident for tasks where a snapshot window may not be enough for detection, such as sepsis (Zhang et al., 2026) and seizure (Tatum et al., 2022) detection. This suggests that how longitudinal window-level representations are modeled can be as important as, or even more important than, the window-level feature extractor itself, which is the main motivation of `BioLT`.

#### Alarm-based evaluation

Figure 6 reports Sens@1FA/h at detection (Section 4.3). `BioLT` recovers the largest fraction of events (0.314), ahead of Snapshot (0.287) and Transformer-XL (0.267); MeanPool, MaxPool, and Attention recover fewer than 8%.

**Figure 6:**
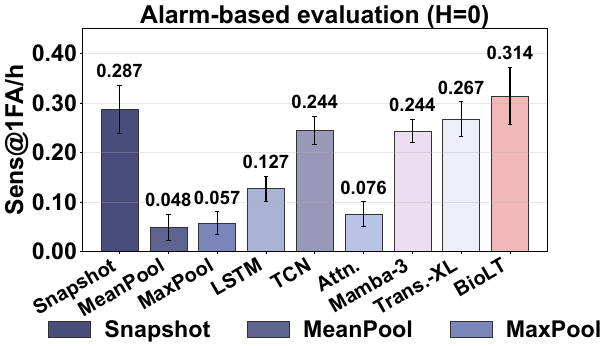
Alarm-based evaluation at detection. (*H*=0). Average Sens@1FA/h over 10 binary dataset settings over 3 seeds.

### 5.2 Performance at Varied Forecasting Settings

#### BioLT remains competitive across forecasting horizons

We next examine how performance changes as the forecasting horizon increases. Figure 7 reports VitalDB ECG binary AF forecasting with CSFM features from detection to 20-minute forecasting. As expected, performance generally decreases as the horizon becomes longer, since the model is asked to predict further into the future from less directly informative observations. Across these horizons, `BioLT` remains consistently competitive with the strongest temporal baselines and often achieves the best or near-best AUROC. This suggests that `BioLT` does not only improve current-state detection but can also support continuously updated prediction over clinically meaningful future horizons.

**Figure 7:**
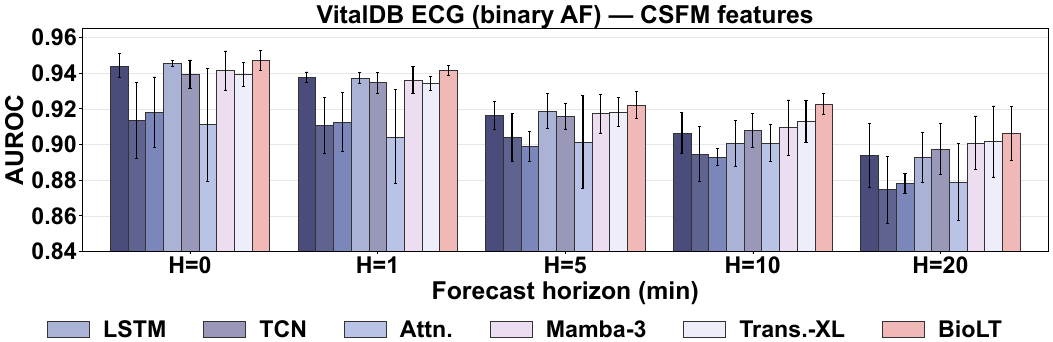
Forecasting across horizons. Forecasting performance on VitalDB ECG binary AF using CSFM features across horizons from *H* = 0 to 20 min. Error bars show the standard deviation over three seeds.

#### BioLT generalizes beyond the fixed-length training context

We further evaluate whether models trained with a maximum context of 500 windows (Section 4.2) can maintain performance when applied to continuously streaming parts of longer recordings. Figure 8 separates test windows into those within the first 500 windows and those beyond this training-context length. Several baselines show variable performance after the 500-window boundary, indicating that extrapolating beyond the training context is non-trivial. In contrast, `BioLT` achieves the strongest performance both before and after the boundary, with no evident degradation beyond the training length. This supports the role of the compressed memory bank in enabling streaming inference over longer recordings.

**Figure 8:**
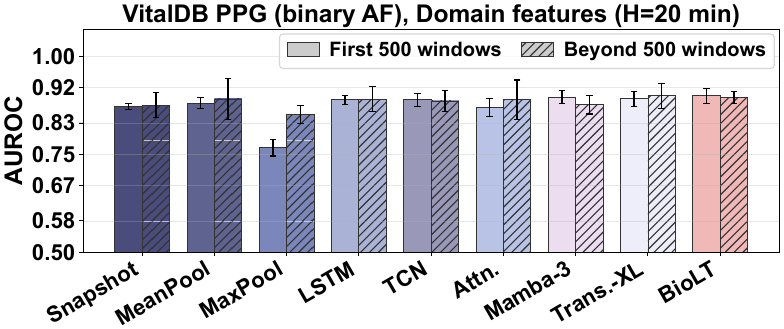
Performance within and beyond the fixed-length training context. Models are trained on at most 500 windows and tested on full recordings; bars show test AUROC within the first 500 windows and beyond, separately.

### 5.3 Ablation Results

#### Validation of compressed memory

To evaluate the “memory” bank actually “remembers” historic information, we keep the backbone frozen, and fit on linear regression layer on top of the captured memory state to reconstruct the mean domain features of chunks located *a* chunks back. This is compared to the memory learned in Transformer-XL and to a Snapshot baseline that sees only the current window. As shown in Figure 9, on VitalDB ECG AF, `BioLT` retains the most information at every distance: it reconstructs the right previous chunk almost perfectly (*R*^2^=0.98, against 0.86 for Transformer-XL and 0.50 for Snapshot) and stays above both up to 120 min, twice the nominal 60-min span of its memory bank.

**Figure 9:**
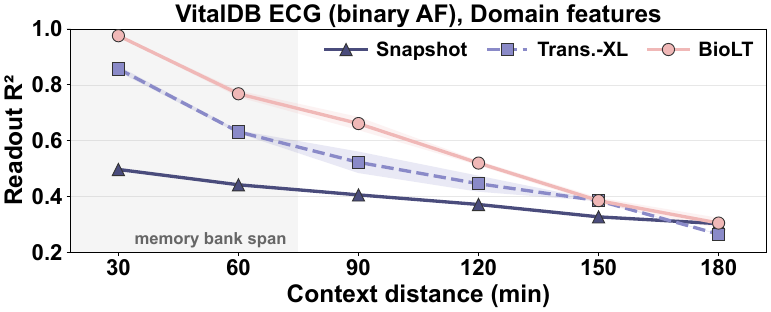
Reconstruction of past context from memory alone. Readout *R*^2^ (mean±std over 3 seeds) of the mean domain features of the chunk located a given distance back. Shaded area: length of the `BioLT` memory bank.

#### Ablation of core components

Furthermore, we ablate two key components of `BioLT`: Δ*s*_*t*_ and memory bank. Full results shown in Table 4 demonstrate the effectiveness of both modules.

**Table 4:** Ablation on MC-MED sepsis forecasting (*H*=10). Mean test AUROC with domain features.

| **Configuration** | **ECG** | **PPG** |
|---|---|---|
| BioLT | **60.29±0.64** | **63.19±0.48** |
| w/o Δ*s* | 58.02±1.71 | 60.61±1.43 |
| w/o mem. bank | 56.68±0.23 | 57.83±0.34 |
| w/o both | 53.82±0.17 | 53.74±0.95 |

## Conclusion

This work studies a practical yet underexplored setting in biosignal learning: how to turn short-window biosignal representations into continuously updated, long-context physiological trajectories. We introduce `BioLT`, a streaming temporal backbone that bridges snapshot representations and continuous clinical monitoring under imperfect, gapped data streams. Across overall 21 dataset-feature settings, covering arrhythmia, sepsis, and seizure tasks, and varied window-level features types, `BioLT` achieves the highest detection AUROC in 15 settings and the highest 20-minute forecasting AUROC in 13. This provides a strong basis on shifting from improving snapshot-level biosignal representations alone toward modelling longitudinal information, which is becoming increasingly accessible through advances in sensing hardware and digital health infrastructure.

## Data Availability

All data analysed in this study are publicly available from PhysioNet (https://physionet.org/), no new data were collected.

https://physionet.org/content/cpsc2021/1.0.0/

https://physionet.org/content/vitaldb/1.0.0/

https://physionet.org/content/mc-med/1.0.0/

https://physionet.org/content/chbmit/1.0.0/

